# COVID-19Predict – Predicting Pandemic Trends

**DOI:** 10.1101/2020.09.09.20191593

**Authors:** Jürgen Bosch, Austin Wilson, Karthik O’Neil, Peter A. Zimmerman

## Abstract

**Background:** Given the global public health importance of the COVID-19 pandemic, data comparisons that predict on-going infection and mortality trends across national, state and county-level administrative jurisdictions are vitally important. We have designed a COVID-19 dashboard with the goal of providing concise sets of summarized data presentations to simplify interpretation of basic statistics and location-specific current and short-term future risks of infection.

**Methods:** We perform continuous collection and analyses of publicly available data accessible through the COVID-19 dashboard hosted at Johns Hopkins University (JHU github). Additionally, we utilize the accumulation of cases and deaths to provide dynamic 7-day short-term predictions on these outcomes across these national, state and county administrative levels.

**Findings:** COVID-19Predict produces 2,100 daily predictions [or calculations] on the state level (50 States x3 models x7 days x2 cases and deaths) and 131,964 (3,142 Counties x3 models x7 days x2 cases and deaths) on the county level. To assess how robust our models have performed in making short-term predictions over the course of the pandemic, we used available case data for all 50 U.S. states spanning the period January 20 - August 16 2020 in a retrospective analysis. Results showed a 3.7% to −0.2% mean error of deviation from the actual case predictions to date.

**Interpretation:** Our transparent methods and admin-level visualizations provide real-time data reporting and forecasts related to on-going COVID-19 transmission allowing viewers (individuals, health care providers, public health practitioners and policy makers) to develop their own perspectives and expectations regarding public life activity decisions.

**Funding:** Financial resources for this study have been provided by Case Western Reserve University.

## Introduction

The pandemic caused by the coronavirus SARS-CoV2, first described from an isolated outbreak in Wuhan, a city of 9.78 million people^1^ first reported in late December 2019, rapidly spread across the globe the genome sequence became available on January 10, 2020^2^. The first reported case of COVID-19 in the United States was on January 20, 2020, in Washington State^3^. The disease caused by SARS-CoV-2, has since been termed Coronavirus Disease 2019 (COVID-19) by the WHO on February 11^4^. As of September 9, 2020 more than 27.8 million cases and 898,426deaths have been confirmed worldwide resulting from COVID-19; the United States alone has 6.3 million confirmed infected and 189,718 deaths attributed to the disease. A general assessment of the global trend of the COVID-19 pandemic, shows that it took 4 months to reach 1 million confirmed cases globally (1.03 M on April 2, 2020) and since then cases have increased by 1 million every 6.16 days (since July 13 worldwide cases have advanced by 1 million every 3.93 days)^4^. Importantly, significant heterogeneity is observed as incidence of new cases varies greatly across different geographic regions. Developing a tool to summarize region-specific changes in COVID-19 cases and deaths is the basis for this study.

In order to track SARS-CoV2 transmission, the Johns Hopkins University COVID-19 Dashboard first emerged with vital data on cases, deaths and recovery^5^. Since then, many more web-based dashboards have appeared from investigative teams around the world emphasizing different perspectives and applying a variety of models to assess the epidemiology of this new disease^6-9^^10^. Results from these efforts have been, and continue to be, critically important in informing federal, state and local response regarding needed supportive facilities and appropriate/necessary equipment and disposable supplies (e.g. intensive care rooms, ventilators, personal protective equipment, nasal swabs, reagents for testing) to support a vast spectrum of COVID-19 medical outcomes^11^. The data used by these dashboards and models have also been used to inform local and regional responses to COVID-19 transmission through recommendations on application of public health measures including social distancing, wearing masks, quarantine and lockdowns of businesses and institutions.

Most of the early modelling forecasts focused on attempts to determine the basic reproductive number (R_o_) and then project long-term (4-24 week) trends of the pandemic without substantial change in transmission. As has been observed, these models have been interpreted in a variety of ways by the general public, national, state and local officials and that actual outcomes often deviate from the long-term predictions. Public health investigators fully expect that community responses to the COVID-19 pandemic will lead to differences between predicted and observed cases and deaths. Furthermore, it is easy to predict that epidemiological outcomes will differ between states, counties, and between different sectors of communities.

Our efforts to develop COVID-19Predict is in response^12^ to turbulence in the representation of the available data across the pandemic timeline for national, state, and county administrative levels by adding an approach to make short-term predictions (7 days in advance) through a continuously, daily updating platform on each of these levels. Through this dynamic process we also examine relationships between testing, morbidity and mortality attributed to COVID-19. In this way, COVID-19Predict may also contribute to unraveling questions pertaining to disease transmission and discovering unknown factors of this disease^13^. At this time, we have restricted our focus to the United States, but our methods enable the same assessments and forecasts for all countries of the world depending on data availability. We anticipate that regular use of the website will be helpful to the general public, clinical and public health care practitioners, and government officials charged with making public health decisions until a safe and effective vaccine emerges and is efficiently distributed to the global at-risk population.

## Methods

### Software and Data Access

Our website is the product of two interacting software components: calculation service and the user interface. An Application Programming Interface (API) was created to integrate the calculation service and the user interface. Both components exist in an Amazon Web Services (AWS) environment. The calculation service runs at 12:00 AM UTC.

Calculations are carried out based on the reported cases and reported deaths from the COVID-19 data sourced from Johns Hopkins University Github. The timeline for the first data point is set to January 21, 2020, the first available data point from the JHU Github server^5^.

### Statistical analysis of historical data

All statistical analysis for this manuscript was carried out using GraphPad Prism 8.4.3.

### Website navigation

Upon arrival at COVID-19Predict, viewers are met with national (admin1) top-tier information. To navigate through COVID-19Predict, progress from top-left to top-right; bottom-left to bottom-right - progressing from Counts to Map to Daily Course of the COVID-19 Pandemic to Accumulated Cases. Each element on the website includes a detailed description by clicking on the “i-information” icon (i) (mobile phone) or by hovering the cursor over a state or county (computer). In the Map panel, the “Map Type” pull-down menu allows for selecting among seven different maps that summarize infection rate, apparent case fatality rate, incidence over the last 14 days, total cases and deaths per 100K population, as well as total predicted cases and deaths per 100K population in the approaching 7 days. Hovering over any state displays the current state-specific counts data – color coding within maps is presented in Table 1. By clicking on any individual state viewers are directed to state (admin2) second-tier information. There, all 4 panels of the display change to report the COVID-19 information for the selected state. In the Map panel, the map of the state appears with all counties or parishes outlined. Hovering the cursor over any county displays the county-specific counts (admin3) as well as the county-specific infection rate and apparent case fatality rate, incidence over the past 14 days, total cases and deaths per 100K population, and total predicted cases and deaths per 100K population in the approaching 7 days. To examine the data for a different state, click “Dashboard” at the top of the page to return to the national level (admin1).

**Table 1.**
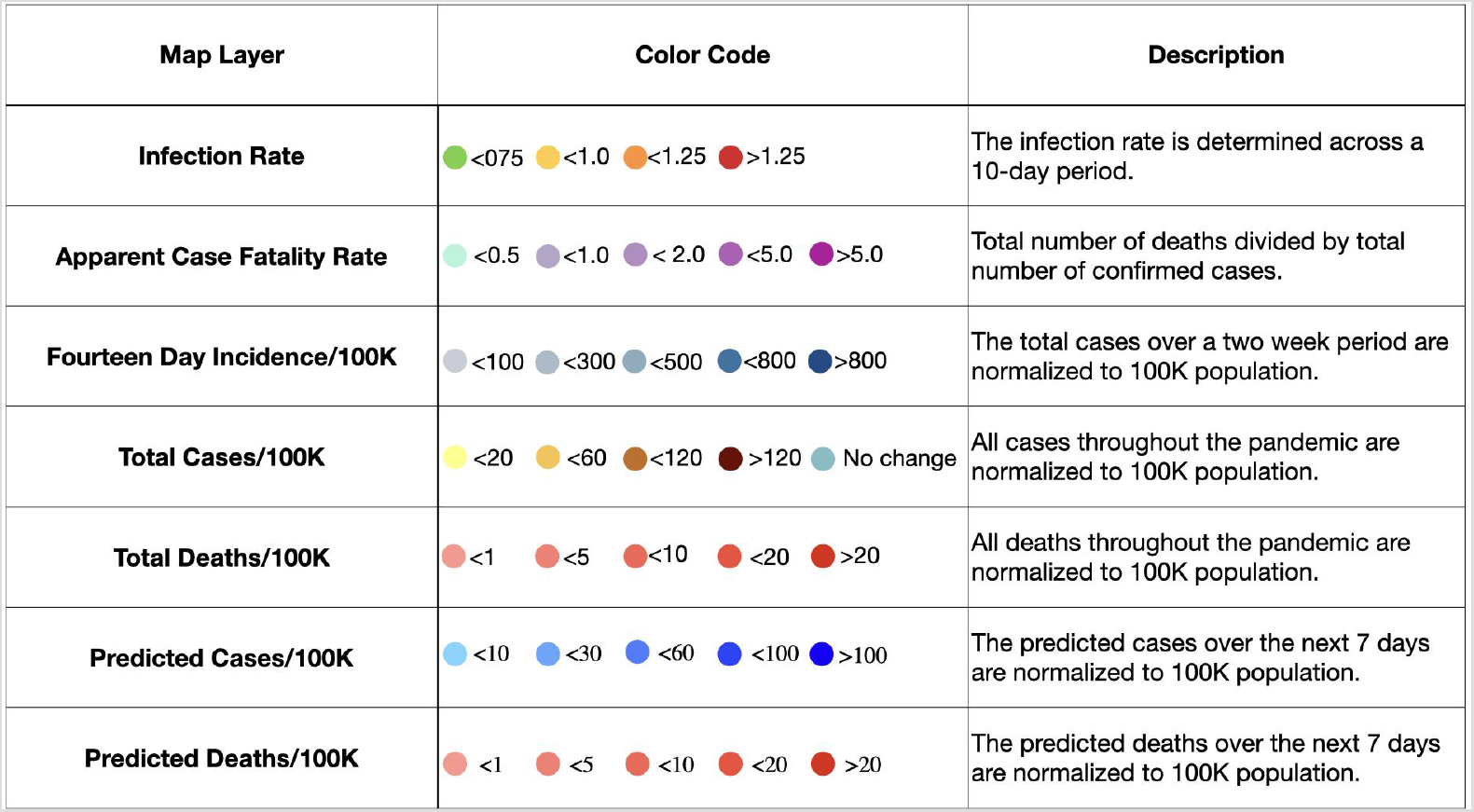
Legends underlying each of the seven map layers, summary counts and statistics

### Daily Course of the COVID-19 Pandemic

Daily cases and deaths are shown graphically as color-consistent flags display the case count (dark blue), death count (red) and date (white). Additionally, the infection rate (yellow) and apparent case fatality rate (magenta) are presented daily. Sliding the diamonds along the bar above the graph allows the user to zoom in for closer examination of specific date ranges.

### Accumulated Cases

In addition to displaying the accumulated cases and deaths graphically, moving the cursor over the graph reveals color-consistent flags displaying the case count (dark blue), death count (red) and date (white). At the right-hand end of the graph, predicted cases and deaths are shown for the next 7 days. Users may zoom in for examination of specific date ranges as indicated above. Clicking on the blue circle with the “-” sign opens the graph to cover the time since the first case was reported in the USA (January 21, 2020).

### Data Analyses

The following calculations generate the data summaries in the map layers and graphs shown in COVID-19Predict (presented in the Results and Discussion section).

### Infection Rate

The infection rate (IR) is determined through a comparison of the confirmed COVID-19 cases across the current 10-day trend in the SARS-CoV2 infection rate.

Infection Rate (IR) is determined by the following equation.

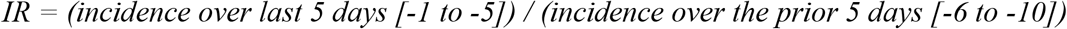

### Apparent Case Fatality Rate

We calculate an apparent case fatality rate (aCFR) as the cumulative number of COVID-19 deaths per the reported, confirmed cases. The aCFR (purple, right-hand Y-axis, logarithmic scale) is displayed as a daily percentage.

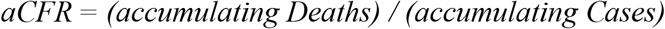

### Fourteen-day incidence (FDI) per 100,000

Assessments compare the Fourteen-day incidence rate per 100K, displaying changes over a larger time period. To compare state and county IRs, we have employed the cutoffs used by the European Center of Disease Control^14^.

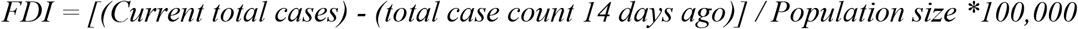

### Weekly Incidence and Death Rates

For the weekly incidence rate per 100,000 population for each calendar week the number of accumulated new cases is normalized to the population of a state or county. The weekly death rate calculation is computed per 100K population as well. Population sizes of states and counties were obtained from Worldometer and hardcoded into COVID-19Predict /our website^15^.

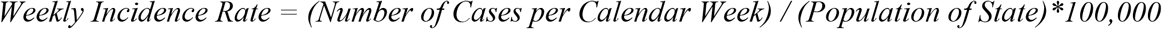

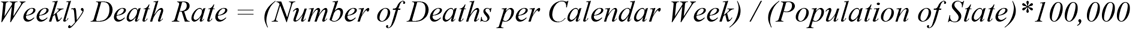

### Total Cases and Deaths per 100,000

All cases and deaths are reported after normalizing to the current 2020 population (national, state, county) accessed through Worldometer^15^.

### Predicted Cases and Deaths per 100,000

The accumulated cases are used to make an exponential curve fit for the last five, seven, and ten days of data to provide an extrapolation for the near-term future. In a fluctuating environment of transmission, illness, and death, the five-day and ten-day fit provide an upper and lower level of confidence for the case and death predictions.

Predicted cases and deaths normalized to the current 2020 population (national, state, county) accessed through Worldometer^15^ were calculated by the following model.

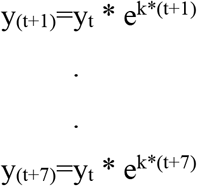

With t = 0 on January 21, 2020 (t = 232 September 8, 2020), y_t_ and k are obtained from fitting either five, seven, or ten days of data to an exponential curve by least squares using the accumulated deaths over the period. Case and death predictions are updated daily and provide estimates for one week into the future for each of the administrative levels evaluated.

## Results and Discussion

All information at the national level (admin1) is the cumulative state level (admin2) and county level (admin3) data Figure 1. These data are displayed on the landing page within four main elements – Counts, Map, Daily Course of the COVID-19 Pandemic, and Accumulated Cases. The map layers available at national and state levels are identical and quantitative information adjusts accordingly (i.e. from national to the specific state) when moving between administrative levels. State and county-level representations are shown at a lower tier in the website with state-specific graphs. Quantitative summaries presented in the graph panels include daily cases, deaths, infection rate (IR) and apparent case fatality rate (aCFR) (lower left); and cumulative cases, deaths and predictive cases and deaths (next 7 days) (lower right).

**Figure 1.**
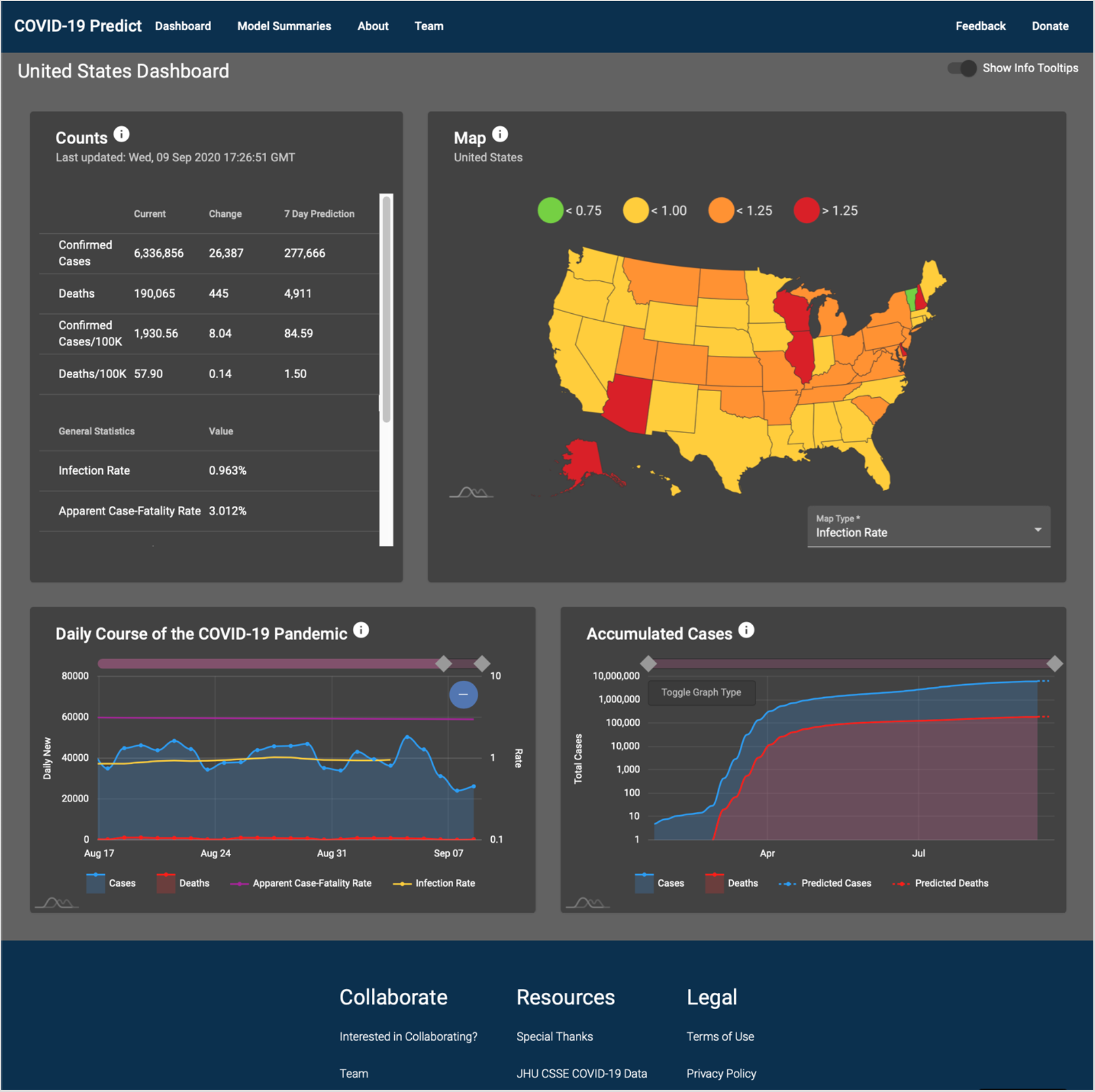
Landing page with key display elements. The landing page summarizes multiple elements of the ongoing COVID-19 epidemic in the United States.

### Critical Evaluation of COVID-19 Epidemiological Data

Reliability or precision of the IR (or similar measures by many other sources) is dependent on the level of testing performed within any individual state. The Covid Tracking Project (https://covidtracking.com/ accumulates and describes this information on a state-by-state basis^16^. From this resource we learn that the number of tests performed varies according to each state (per capita tests from 0.102 to 0.638) and that across the United States, SARS-CoV-2 testing is performed at just 62% of the rate recommended to achieve mitigation. Additionally, enumeration of tests varies from (a) all tested samples counted, regardless of individuals being tested multiple times (6 states plus the District of Columbia); (b) all PCR tested samples counted regardless of multiple sampling (as in “a”) but corrected for antigen testing (34 states plus Puerto Rico); (c) only the first sample collected from a person counted - multiple samples excluded (9 states). This brief summary suggests there are multiple indicators suggesting that COVID-19 testing is performed following a variety of protocols across the United States, unavoidably leading to inconsistencies. This suggests it may be difficult to compare the quantitative benchmarks for testing that epidemiologists use to characterize the novel coronavirus between and within states.

Given a significant potential for uncertainty in these testing benchmarks, we reason that deaths attributed to COVID-19 (assessed through PCR testing, serology, multiple clinical markers and formalized protocol) may be more reliable than case/infection estimates^17-19^. We have therefore calculated the aCFR (magenta, right-hand Y-axis, logarithmic scale) as a proxy to monitor potential inaccuracies in estimating the number of infected individuals. Others have termed this to be a measure of the infection fatality rate (IFR)^20,21^. Both approaches emphasize that estimating the number of infected individuals is made very challenging for COVID-19 tracking because (1) the high number of SARS-CoV-2-infected individuals who are asymptomatic and do not get tested and (2) limitations to diagnostic testing. Overall, we have observed a range in aCFR among the 50 states from 0.718 to 8.45 and a range among the 3,142 counties from 0 to 83.6. We interpret the ratio of the aCFR to represent a multiplier to more accurately predict the number of SARSCoV-2-infected individuals in an administrative level when compared with more broadly calculated CFRs (current CFRs calculated by the United States CDC = 0.68%, median international CFR - 0.27%)^20,22^. Therefore, if the current aCFR is 6.8%, this would be 10-fold higher than the CDC calculated CFR and 25.1-fold higher than the international CFR. This may suggest that given 1,000 reported cases within the admin level the actual number of cases could be between 10,000 and 24,000. Alternatively, a local aCFR that is higher than expected, could signal the emergence of new, more serious cases or indicate that a local health care delivery system was failing its patient population.

Further evaluation of the data, the analytical summaries, and the 7-day forecasting methods has been performed in developing COVID-19Predict to assess the variability of the predictions and observed outcomes (Figure 2, Supplemental Figures S1, Supplemental Table 1). To do so we performed identical calculations as outlined previously for five, seven, and ten day intervals moving forward one day at a time starting five, seven or ten days after the first case was reported in all states through August 2020. Results of this analysis presented in Figure 2A show that the percent difference between the observed data and 7-day prediction outcomes decreased significantly from the 5-day to 10-day approaches (3.7% for 5-day; 1.1% for 7-day; −0.2% for 10-day data). The same data are shown in Figure 2B for all states corresponding to the state’s population size; inset graph indicates the range in the number of observation days since the beginning of the COVID-19 epidemic in the United States (mean days = 162.2). The assessment in Figure 2B also suggests that while smaller states may show greater variability in predicting SARS-CoV-2 infection trends, the 7-day predictions and data comparisons have also varied for some larger states.

**Figure 2.**
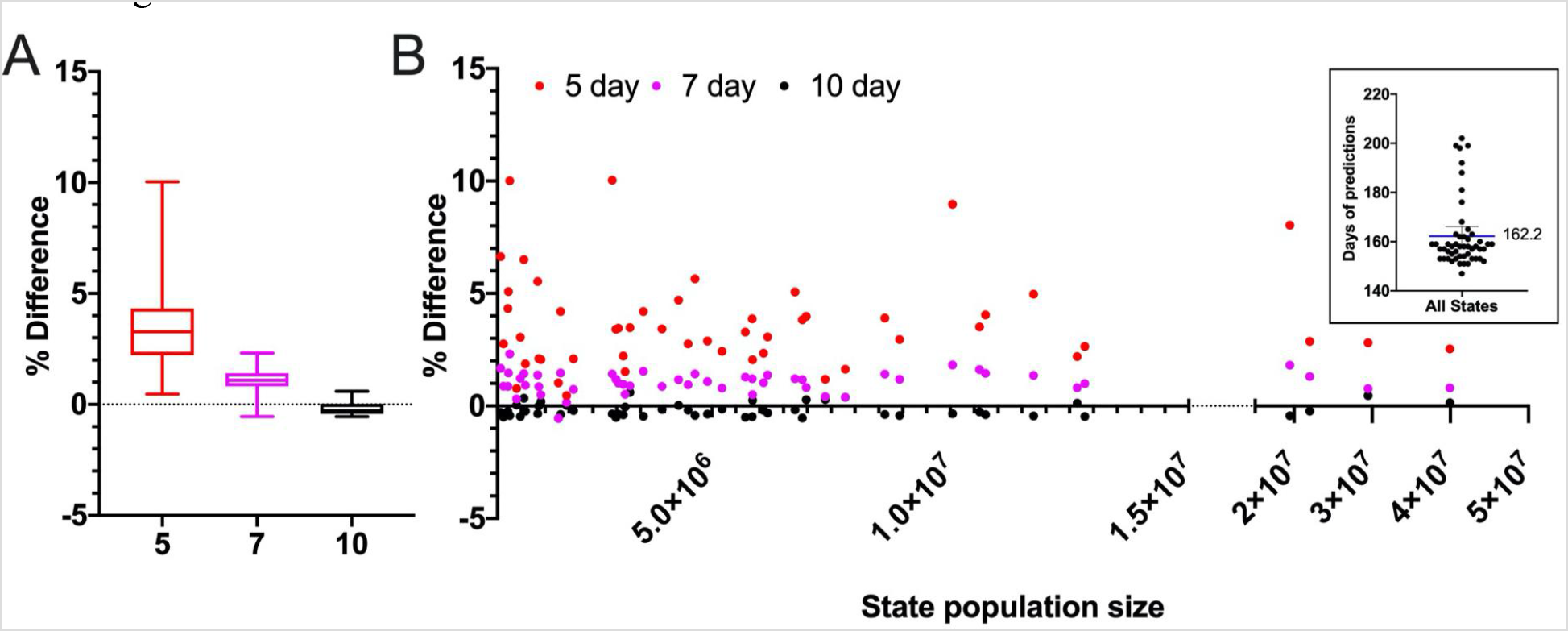
Assessment of 7-day prediction model over the course of the pandemic. A) The graph shows the mean percent difference between actual and predicted cases per state using either five (red), seven (magenta) and ten (black) days to predict 7 days into the future. B) The graph displays the percentage difference between prediction and actual case count versus the population for all 50 US states. The inset shows the number of predicted days per state plotted (mean = 162.2), with Washington (first state where COVID-19 was reported) having the highest number of days (n = 202) and West Virginia (last state where COVID-19 was reported) with the lowest number of days (n = 147). Statistical analysis was performed using GraphPad Prism.

### Comparisons Among COVID-19Predict Map Layers

Further assessments of the predictive value of the daily infection data to predict the approaching trend of COVID-19 cases are presented for each state in Supplemental Figure S1, Supplemental Table 1. Overall, it is consistently observed that the largest discrepancies (between observed and predicted cases) were at the beginning of the pandemic when few cases were present. Supplemental Table 1 reports that the average percentage difference between observed and predicted case counts was 3.69% using 5-day prior data (range of lower and upper 95% CI of the mean: 0.46 to 10.03%), 1.06% using 7-day prior data (range of lower and upper 95% CI of the mean: −0.55 to 2.31%) and –0.19% using 10-day prior data (range of lower and upper 95% CI of the mean: −0.55 to 0.60%). Overall the accuracy increases when more days are used to fit the exponential growth curve. This held true irrespective of the state population size as observed in California (largest population size of 39.9 million; predictions based on 5-day = 2.54% mean difference, 7-day = 0.8%, 10-day = 0.14%) and in Wyoming (smallest population size of 567,025; predictions based on 5-day = 6.64% mean difference, 7-day = 1.67%, 10-day- −0.29%). Therefore, while there are many differences in the way COVID-19 is diagnosed across the United States (as discussed above), the trends analyzed are shown to lead to reproducible predictions of on-going COVID-19 infection.

**Figure 3.**
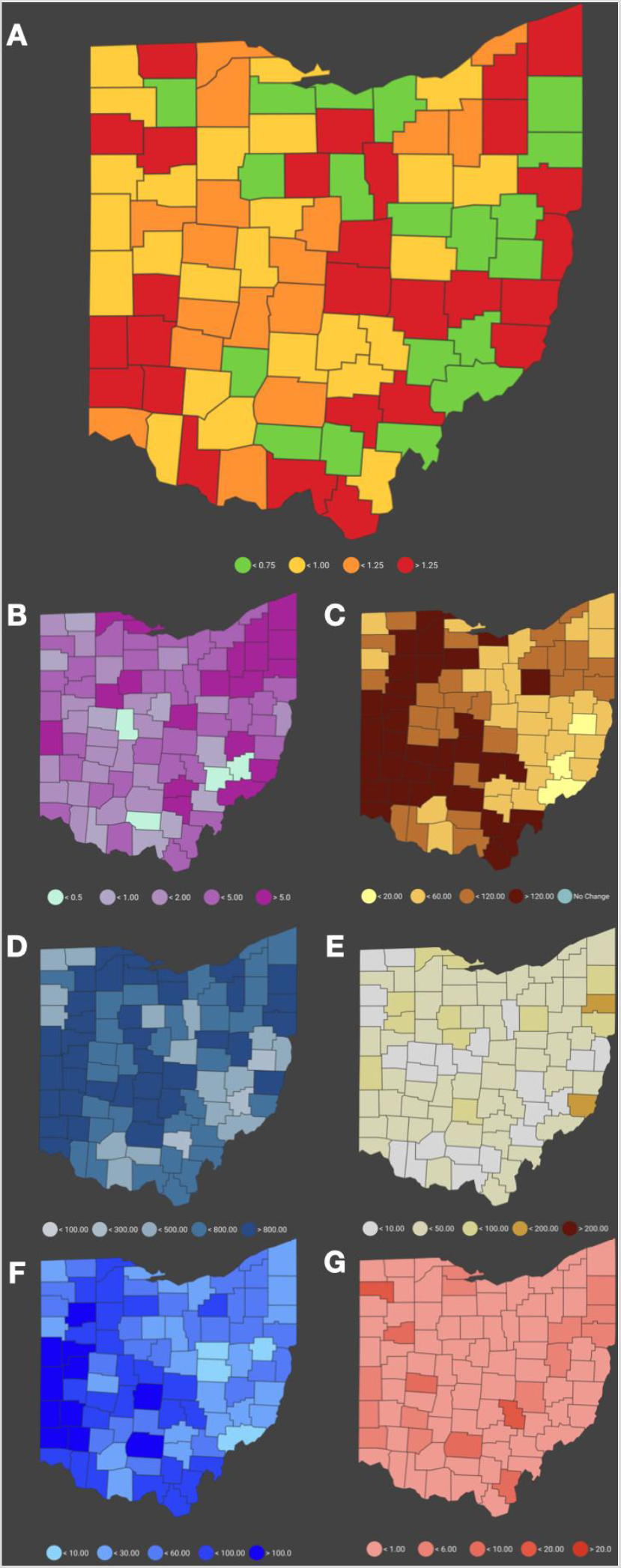
Covid19Predict Map layers. A) Infection Rate (IR), calculated over the past ten days. B) The apparent Case Fatality Rate (aCFR) calculated over the whole period of the pandemic. C) Fourteen-day incidence (FDI) rate. D) Cases per 100K. E) Deaths per 100K. F) The 7-day prediction of cases is visualized per 100K population. G) The 7-day prediction of deaths is visualized per 100K population. Legends to interpret quantitative differences across counties are provided below each map and are consistent with Table 1.

Map layers displayed in Figure 3 present multiple facets of infections and deaths attributed to COVID-19 including current characteristics (3A to 3C; IR, aCFR and FDI per 100,000, respectively), cumulative characteristics (3D and 3E; total cases and total deaths per 100,000, respectively) and predicted characteristics, 7 days into the future (3F and 3G; predicted cases and total deaths per 100,000, respectively); map types and order are identical for national and state maps. The state IR map provides insight into how local county transmission is contributing to the state outcome. Further, the aCFR map allows an assessment of the potential severity of disease, stress on the local health care system, and/or the level of under-testing in a county. For example, when a county shows a decrease in the IR but the aCFR is high, this comparison would suggest caution and that the actual number of cases in the county is likely to be higher by the aCFR multiplier. Map layers 3C to 3G are presented per capita to facilitate comparisons between counties or states based. The FDI/100,000 and Total Cases/100,000 offer broader perspectives on COVID-19 incidence over longer time-frames providing additional points of references for comparison with the more rapidly changing IR, with the FDI/100,000 providing an intermediate perspective (Figures 3C and 3D). This intermediate perspective is useful given that the Total Cases/100,000 categorical coloration (introduced by the Robert Koch Institute (Berlin) and used in European countries) is over 800/100,000 in 2,210 of the 3,142 U.S. counties (68.5%). Where overall incidence is lower, similarities between these measures are observed, as in western Ohio. Additionally, these comparisons can reveal areas where notable decreases in COVID-19 incidence has occurred (e.g. Mid-Hudson, New York City and Long Island regions). The comparisons in total cases and deaths per 100,000 clearly show the accumulation of disease burden across the United States (Figures 3D and 3E). However, accumulation of cases beyond 800/100,000 is not highly correlated with deaths per 100,000. Finally, the comparisons between predicted cases and deaths per 100,000 provides a perspective on where pending stress to local health systems is most likely to be observed (Figures 3F and 3G). These predictions may be particularly important to rural communities in which resources to provide medical care and treatment are limited and public health programs have been impacted through decreased funding.

### Dynamic Changes in COVID-19 IR

In calculating the IR, a 5-day interval was chosen as the average person-to-person transmission time for SARS-CoV2 is 5.2 days^23^. On the county level if less than 100 cases per 5-day interval are reported, the infection rate becomes less accurate. Since the IR is back-calculated, the last IR is displayed 5 days prior to the current date (yellow, right-hand Y-axis, logarithmic scale). The infection rate is not to be confused with the basic reproduction number R_0_, which describes how many resulting infections a single carrier can cause (current best estimate provided by the CDC for the USA COVID-19 R_0_ = 2.5)^24^. Variability in the IR from state-to-state is reflected in the varying color pattern in the national map and from county-to-county in the state maps. These patterns capture the dynamic nature of COVID-19 incidence across the different administrative levels. Across all geographic landscapes the goal of this predictive model is to assist the public in reducing the IR below rates where every single infection is predicted to lead to infection of one or more uninfected individuals. Therefore, the green and yellow IR categories are the more positive indicators that SARS-CoV-2 transmission is being constrained by public health measures, while the orange and red categories indicate that transmission is expanding the number of infected people.

A demonstration of the dynamic variation in IR is provided below in Figure 4. The states of California, Connecticut, Ohio and Vermont have been selected to show the daily change in IR across counties within the states and the impact these local dynamics have on the state IRs in the national map between August 16, 2020 to August 22, 2020 (Top to bottom). During this time, California decreased the number of counties with increasing IRs (red and orange) from 43 of 58 (74.1%) to 21 of 58 (36.2%), transitioning state-wide from red (>1.25) to yellow (< 1.0). Connecticut transitioned from red to yellow and back to red across this time, as 7 of 8 (87.5%) counties experienced increasing transmission at the beginning of the time period, 6 of 8 (75%) experienced decreasing transmission in the middle and there were equal numbers of counties with increasing and decreasing transmission 4 of 8 (50.0%) as of August 22. Across Ohio (transitioned from orange to yellow (6 of 7 days), the majority of the state’s 88 counties showed evidence of decreasing transmission (54.5% to 70.5%). This observation window occurred at the end of a significant reduction in SARS-CoV-2 transmission from just after the July 4th holiday (daily cases = 1,378) through August 10th (daily cases = 613). While the majority of the counties in Vermont consistently showed evidence of decreasing transmission, Burlington (the state’s most populous city and having the greatest number of COVID-19 cases) resides in Chittenden county (only red county in 6 of 7 time points). As a result, Vermont was consistently shown to have an increasing IR until Chittenden transitioned to yellow on August 22.

**Figure 4.**
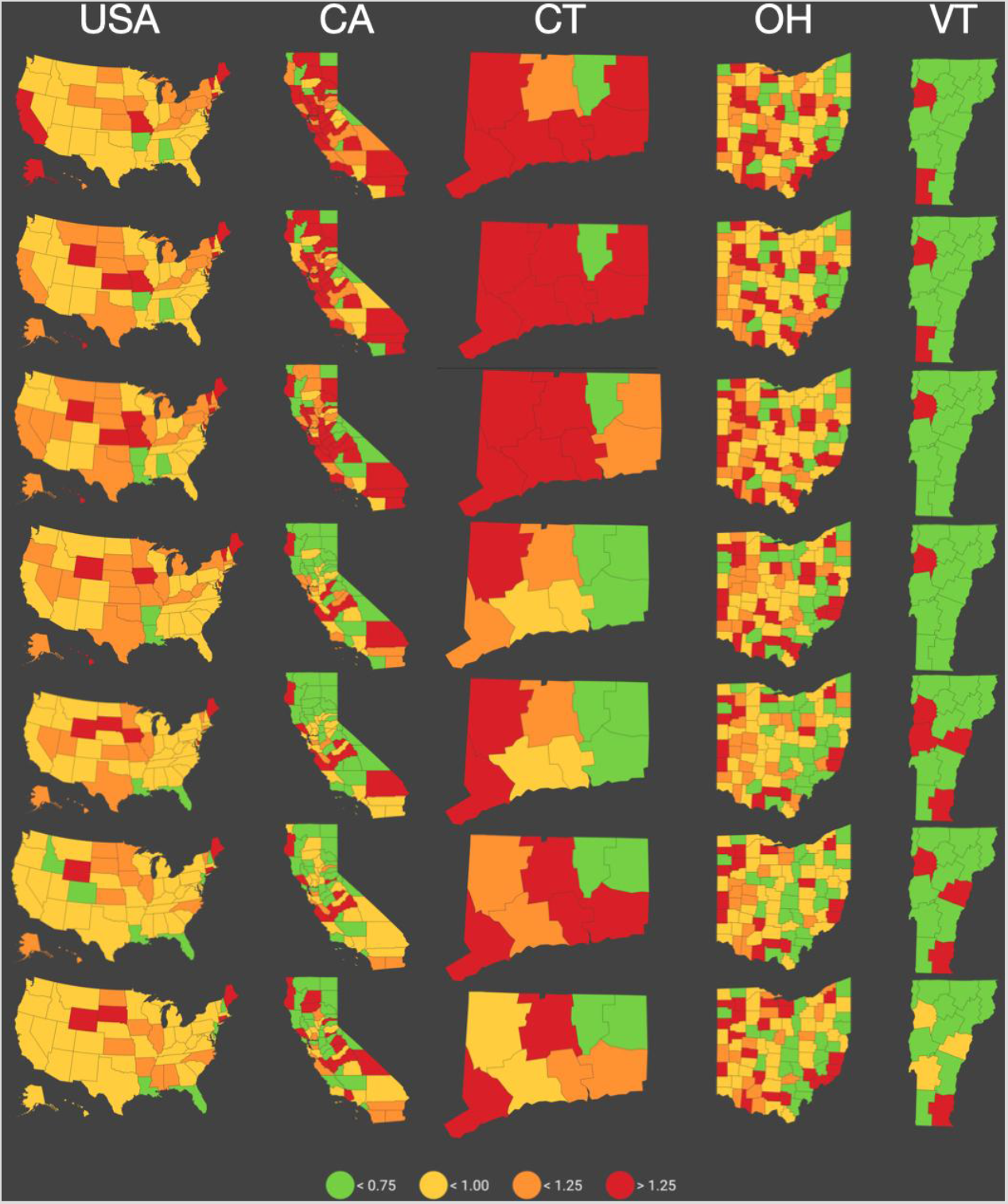
Longitudinal observations of COVID-19 incidence for states with varying infection and mortality rates. Accumulating data as of September 6, 2020 in selected states cover the spectrum of COVID-19 cases, deaths, infection rates and apparent Case Fatality Rates (California (CA) = 736,209 cases, 13,709 deaths, IR = 0.903%, aCFR = 1.9%; Connecticut (CT) = 53,356 cases, 4,468 deaths, IR = 1.108%, aCFR = 8.9%; Ohio (OH) = 129,785 cases, 4,256 deaths, IR = 1.117%, aCFR = 3.3%; Vermont (VT) = 1,647 cases, 58 deaths, IR = 0.977%, aCFR = 3.5%.

## Conclusion

Using publicly available data from the JHU Github^5^, we developed COVID-19Predict as a tool to guide interested users through the continuing SARS-CoV-2 pandemic. The daily predictions made by COVID-19Predict down to the county level provide useful insights with respect to potential risks for infection and help guide individuals’ activities and community planning. Retrospective assessments make it easy to observe how increased social behaviors around the Memorial Day and July 4th holidays contributed to increased infection, followed by increased deaths in many parts of the country. With the reopening of schools across the country, further dynamic changes presented by COVID-19Predict can be expected to detect increases in local transmission. At the launch of this website we have focused on making our predictions at the county, state and national level for the United States. Given the ever-changing dynamics of the pandemic and that country-to-country and continent-to-continent travel and commerce are influencing and influenced by continuing outbreaks, we are in the process of expanding the analyses performed here to all countries where data is available. This perspective will help facilitate learning what measures have helped to constrain COVID-19 and how these public health strategies can begin to reduce the public health threat of this disease.

## Limitations

COVID-19Predict relies on the data reported by hospitals to the JHU server to be an accurate representation of the current status of SARS-CoV-2 transmission and COVID-19 morbidity and mortality. If there are disruptions to the availability of this data, the presentations provided in COVID-19Predict and many other COVID-19 dashboards will be compromised.

## Data Availability

Data will be made available upon request.

https://covid19predict.com/dashboard

## Contributors

JB, AW, and PAZ contributed to study concept and design. JB, AW contributed to acquisition of data. JB, AW, KO contributed to data analysis. JB and PAZ contributed to initial drafting of the manuscript. All authors contributed to interpretation of data and critical revision of the report. JB and PAZ contributed to study supervision.

## Acknowledgements

The authors acknowledge the use of the data provided by Johns Hopkins Center for Systems Science and Engineering on their COVID-19 Dashboard and the underlying Github. Presentation of the measurements of health indicators herein are consistent with the Guidelines for Accurate and Transparent Health Estimates Reporting (GATHER)^25^. We thank Drs. Karen Abbott, Peter Thomas and Michael Wilson for critical review of the manuscript and our numerous beta-testers of COVID-19Predict for their candid feedback leading to implementation of additional features and improvements.

## Human Participant Protection

This work was deemed to not be human participant research and was thus exempt from protocol review.

